# More spice, less salt: how capsaicin affects liking for and perceived saltiness of foods in people with smell loss

**DOI:** 10.1101/2023.06.05.23290966

**Authors:** Stephanie R. Hunter, Candelaria Beatty, Pamela H. Dalton

## Abstract

People who lose their sense of smell self-report consuming more salt to compensate for a lack of flavor and enhance eating enjoyment. However, this can contribute to excess sodium intake and a poor diet. Capsaicin may help increase salt taste intensity and eating enjoyment in this population, but this has not been studied. The purpose of this study was to determine 1) whether salt intake in those with smell loss differs from population averages, 2) whether capsaicin increases flavor and salt taste intensity, and 3) if adding spice to foods increases food liking in individuals with smell loss. Participants 18-65 years old with confirmed partial or total smell loss for at least 12 weeks completed two sets of replicate test sessions (four total). In two sessions participants rated overall flavor intensity, taste qualities’ intensities, spicy intensity, and liking for model tomato soups with low or regular sodium content and three levels of capsaicin (none, low, or moderate). In the other two sessions, participants rated the same sensory attributes for model food samples with three levels of added spice (none, low, or moderate). 24-hour urine samples were also collected to determine sodium intake. Results indicate that although sodium intake is higher than recommended in those with smell loss (2893 ± 258 mg/day), they do not consume more sodium than population averages. Adding low and moderate amounts of capsaicin to a model tomato soup increased the intensity of overall flavor and saltiness compared to a model tomato soup without capsaicin. However, the effect of capsaicin on liking differed by food type. In conclusion, the addition of capsaicin can improve flavor, salt taste intensity, and eating enjoyment in people with smell loss.

## 1. Introduction

Prior to the COVID-19 pandemic, it is estimated that about 20% of the population suffered from long-term smell disorders^1–4^. Since the COVID-19 pandemic began, in which smell loss is a cardinal symptom^5^, millions more people have lost their sense of smell long-term^6,7^. Smell disorders are more prevalent now than ever before, and more people than ever will suffer from their negative effects on physical and mental health.

Food odors contribute to flavor and a pleasurable eating experience^8,9^. One of the primary complaints in people who lose their sense of smell is the lack of flavor perception and reduced eating enjoyment^10,11^, which negatively affects their quality of life^12,13^. People with smell loss often report compensatory strategies in attempts to regain flavor and increase palatability of foods. One of these compensatory strategies is adding more salt to foods. Those with smell loss self-report being more frequent salt consumers^11,14^ and preferring salty foods after losing their sense of smell^10^. However, if this is maintained with long-term smell loss, this could lead to excess sodium intake. Another compensatory strategy that people with smell loss often report is adding hot sauce or spiciness to foods, which increases the chemesthetic aspect of flavor^10,11,15,16^ . For example, one person with smell loss stated that “…I opted for more spicy food… stuff with a lot more heat in it just to taste something”^15^. Thus, this may be a useful strategy to improve flavor and palatability, but this has not been tested.

Sodium intake is already too high throughout the population. The World Health Organization recommends consuming 5 grams of sodium per day, but the average intake is 9-12 grams per day^17^. In the United States, the dietary guidelines recommend consuming less than 2300 mg of sodium per day^18^, however the average sodium intake is 3,361 mg/day^19^. Many health organizations have goals to reduce sodium intake^18,20–22^. If those with long-term smell loss are consuming even more sodium after losing their sense of smell, this could exacerbate the problem that already exists, and hinder population wide efforts to reduce sodium intake. Compensatory strategies that improve flavor, but do not negatively affect health should be encouraged in those with smell loss^23^.

Previous studies have found that capsaicin can increase salt taste intensity in people with a normal sense of smell. In one study, normosmic individuals reported that 4.4 g/L solutions of salt tastes saltier with 0.1 or 0.3 mg/L added capsaicin compared to when no capsaicin is added^24^. Others have found that 3200 mg/L of capsaicin increased salt taste intensity of a 0.98 g/L salt water solution by 51% compared to the salt water solution with no capsaicin added^25^. Observational studies also report that participants with a high spice preference had lower salt intakes and blood pressure than individuals who disliked spicy food^26^. Given that individuals with smell loss self-report adding spicy ingredients onto foods to regain some flavor, adding a small amount of capsaicin to foods may be a well-adhered to strategy to improve flavor and food liking while avoiding excessive salt intake in these individuals. However, this strategy has not been tested in individuals with smell loss.

The purpose of this study was to 1) establish current sodium intake and eating behavior changes in those with smell loss - we hypothesized that there will be differences in eating behavior after smell loss and that current sodium intake will be high compared to recommended levels; 2) determine if capsaicin increases salt taste sensitivity in individuals with smell loss - we hypothesized that the addition of capsaicin to an NaCl solution will increase salt taste sensitivity; and 3) quantify liking of a meal with and without capsaicin in individuals with smell loss - we hypothesized that adding capsaicin to a food in those with smell loss will increase liking compared to the same food without capsaicin.

## 2. Methods

### 2.1 Participants

Participants were recruited from the greater Philadelphia, PA area from October 2021 to September 2022. Eligibility criteria included men and women 18 - 65 years old with acquired anosmia or hyposmia for at least 12 weeks. Smell loss was confirmed using the NIH Toolbox Odor Identification Test^27^ and a SCENTinel^TM^ test^28^. Those who answered 6 or less questions correctly on the NIH Toolbox Odor Identification Test and/or rated the odor intensity on the SCENTInel_TM28_ test ≤ 50 (out of 100) were included in this study. Those who had congenital anosmia were excluded from this study due to potential differences in eating behaviors from those with acquired smell disorders^29^.

33 participants were included in this study (Table 1). Results from the NIH Toolbox Odor Identification Test indicated that 10 had anosmia, and 23 had hyposmia. Furthermore, 11 of those with hyposmia also self-reported that they experienced parosmia. 88% of participants were female, 85% were white, and the average age was 47 ± 11 years old. The average smell loss duration was 3 ± 5 years, from a variety of etiologies, including COVID-19, surgery, traumatic brain injury, and unknown reasons.

**Table 1.** Demographics.

| Characteristic | Participants |
| --- | --- |
| n | 33 |
| Age; (mean ± SE) | $47 \pm 11$ |
| Sex; n (%) | 29 (88%) Female<br>4 (12%) Male |
| Race; n (%) | 28 (85%) White<br>3 (9%) Black<br>2 (6%) Mixed Race |
| Objective Smell Disorder; n (%) | 10 (30%) with Anosmia<br>23 (70%) with Hyposmia |
| Length of Smell Disorder; (mean ± SD) | $3 \pm 5$ years |
| Frequency of Spicy Food Consumption; n (%) | 2 (6%) never or less than 1 time per month<br>1 (3%) 1 time per month<br>9 (27%) 2 - 3 times per month<br>2 (6%) 1 time per week<br>10 (30%) 2 - 3 times per week<br>3 (9%) 4 - 6 times per week<br>1 (3%) 1 time per day<br>5 (15%) Unknown or not reported |

### 2.2 Design

This was a within-subjects study. Participants came to the Monell Chemical Senses Center for four, 1-hour test sessions, which consisted of two replicate test sessions: two in which they completed sensory evaluations of model tomato soups, and two in which they completed sensory evaluations of two other model foods (pasta and chocolate). Order of the sessions was counterbalanced across participants, such that about half were randomly assigned to sensory evaluations of model tomato soups first, and model foods second, and others were randomly assigned to sensory evaluations of model foods first, and model soups second.

### 2.3 Procedure

This research was performed in accordance with the Declaration of Helsinki and was approved by the University of Pennsylvania Institutional Review Board (protocol #849791). All participants provided written informed consent prior to taking part in this study.

Participants arrived fasted for at least one hour prior to each test session. For one of the replicate test sessions, participants first completed scale training. Participants were then provided with 12, 30-mL samples of tomato soup. In a random order, participants swished the sample around their mouth, and swallowed the sample. They then made intensity and liking ratings on the sample. Participants waited for two minutes in between samples, cleansed their palate with a cracker (Nabisco Saltine Cracker, Unsalted Tops), rinsed their mouth with whole milk^30^ (SKU 8826700874, Foodhold USA, LLC), and expectorated the milk. For the other replicate test sessions, participants first completed scale training. Next, participants were provided with six, 30-g samples of pasta, followed by six, 15 g samples of chocolate. For each food, participants tasted the sample, swallowed it, and then made intensity and liking ratings. Participants waited 2 minutes in between samples, cleansed their palate with a cracker (Nabisco Saltine Cracker, Unsalted Tops), rinsed their mouth with whole milk (Foodhold USA, LLC), and expectorated the milk. At the end of the first test session, participants completed a questionnaire to determine how their eating behaviors differ from before versus after losing their sense of smell, adapted from Aschenbrenner et al^10^. They were also provided with supplies to collect their urine for a 24-hour period.

### 2.4 Stimulus Materials

Tomato soup samples were prepared using commercially available Regular Sodium (Imagine Foods Creamy Tomato Soup; 670 mg Sodium) and Reduced Sodium (Imagine Foods Light in Sodium Garden Tomato Soup; 280 mg Sodium) soup. Based on methods from Narakuwa et al^24^, 0, 0.0003, or 0.0006 g of capsaicin was added per liter of soup, and from now on will be referred to as none, low, or moderate amounts of capsaicin, respectively. Samples were coded with three digits numbers, and there were different codes for replicates. Participants were provided each sample in a random order, and in duplicate, such that they sampled 12 tomato soup samples during the respective test session. Soups were served at 150°F and kept hot using a water bath.

Pasta and chocolate samples were used to simulate a more meal-like experience. Pasta samples were prepared according to the Sambal butter noodles recipe from the Taste & Flavour Cookbook by Ryan Riley and Kimberley Duke, which was created specifically for individuals who lost their sense of smell because of COVID-19^31^. Specifically, 260 g of pasta (Light ‘n Fluffy extra wide egg noodles, SKU 3340061280) was cooked for 6 minutes, and then strained. Seventy-five grams of unsalted butter (SKU 8826707471, Foodhold USA, LLC) was added to the pasta, along with 2 tablespoons of dried basil (SKU 8826701570, Ahold USA, Inc.), 100 g of parmesan cheese (SKU 8826716875, Ahold USA, Inc.), ¼ teaspoon of salt, and ¼ teaspoon of pepper, and mixed well so that each noodle had an even coating of ingredients. One-hundred grams of the pasta was divided into three different bowls. One bowl had no sambal oelek (Huy Fong Foods, Irwindale, CA) added (no spice), one had ¼ teaspoon of sambal oelek added and mixed well (low spice) and had ½ teaspoon of sambal oelek added and mixed well (moderate spice). Thirty grams of each pasta was served to participants in duplicate in clear, plastic souffle cups, coded with a three-digit number. Replicates were coded with different three-digit numbers. Chocolate was prepared by melting 73 g of chocolate (Hershey’s Milk Chocolate Baking Chips) in a double broiler until it reached 110-115°F, removing it from heat and cooling to 95-100°F, then adding in an additional 36 g of chocolate, and stirring until melted. Then, either 0g, 0.17g, or 0.33g of cayenne pepper was added and mixed thoroughly. The chocolate was poured into a mold and cooled completely. Fifteen grams of each chocolate was served to participants in duplicate in clear, plastic souffle cups, coded with a three-digit number.

### 2.5 Scale Training

At the beginning of each test session, participants were trained to use the general labeled magnitude scale (gLMS)^32^ and the labeled hedonic scale (LHS)^33^ using standard instructions, then practiced each scale by rating the intensity or liking of imagined sensations. The gLMS is a vertical scale with intensity descriptors of “barely detectable”, “weak”, “moderate”, “strong”, “very strong”, and “strongest imaginable sensation of any kind”, with the spacing of the descriptors on the scale determined empirically to be proportional to the strength of the sensation. The LHS is a vertical scale with liking descriptors of “most disliked sensation imaginable”, “dislike extremely”, “dislike very much”, “dislike moderately”, “dislike slightly”, “neutral”, “like slightly”, “like moderately”, “like very much”, “like extremely”, and “most liked sensation imaginable”.

### 2.6 Ratings of intensity and liking

For each sample, participants rated the intensity of flavor, taste qualities (sweet, sour, salty, bitter, and umami), and spiciness using a gLMS, and how much they liked the sample using a LHS, in that order. Umami was described to participants as how savory the sample was. While we were specifically interested in salt taste intensity, we assessed all taste qualities to prevent a dumping effect^34^.

### 2.7 Dietary Behaviors Questionnaire

Changes in dietary behaviors before and after smell loss were assessed using the Dietary Behaviors Questionnaires, as done in Aschenbrenner et al 2008^10^. This is a 41-item questionnaire with questions about taste preferences, body weight changes, social eating behaviors, and whether different foods and nutrients are consumed more, less, or no change now compared to before losing their sense of smell. Two questions to determine changes in salt intake (“Do you use more or less salt now?”) and spicy intake (“Do you eat more or less spicy foods now?”) were added to the survey.

## 2.8 24-hour urine collection

Participants were provided with supplies to collect their urine for a 24-hour period after the first test session. Participants were instructed to void their first urine upon waking up, and then to collect all urine including the final specimen voided at the end of the 24-hour collection period. 24-hour urine volume was recorded, and a 10-mL aliquot was sent to LabCorp to assess sodium content.

### 2.9 Data Analysis

The sample size calculation for this study was based on a power analysis that indicated a sample of 30 participants would be sufficient to detect a moderate effect size of 0.35 with 95% power for detecting differences in rated salt taste intensity. The α level was set at 0·05 for all analyses. To account for potential dropouts, our goal was to recruit 35 participants, but due to time constraints, we stopped data collection after we reached 33 participants. Not all participants completed all study sessions, and the sample size included in each analysis is described below. Data are reported as means and standard errors unless otherwise stated.

All 33 participants completed the Dietary Behaviors Questionnaire. The Dietary Behaviors Questionnaire was analyzed using chi-square tests, Kruskal-Wallis tests, and t-tests where appropriate. 24-hour urine was compared to population averages using t-test, and compared across objective smell disorder groups using one-way ANOVA. Urine volumes less than 500 mL were considered incomplete and were excluded from the analysis^35–37^. One participant had a 24-hour urine volume less than 500 mL. Twenty-five participants had complete 24-hour urine samples and were included in the analysis.

We performed a test-retest analysis on all ratings for replicate samples within and between sessions. The test-retest reliability ranged from 0.16 – 0.7, and all ratings were significantly correlated within and between sessions (see Supplementary Tables 2-5). Thus, we averaged ratings across replicate samples using arithmetic means for the analyses. Linear mixed models were used to determine the effect of sodium (regular or reduced), capsaicin (none, low, moderate), and sodium*capsaicin [for soup samples; n=29], or food (pasta or chocolate), spice (none, low, or moderate), and food*spice [for food samples; n=31] on total flavor, taste qualities, and spiciness intensities, and liking. Sodium and capsaicin, and food and spice levels were fixed effects, and participants were treated as random effects. Age, sex (male, female, other, prefer not to answer), frequency of spice consumption, length of smell disorder, and objective smell disorders were included in the models as covariates. When the main or interaction effects were significant, pairwise comparisons were conducted with Bonferroni correction for multiple comparisons. Variables that were not Normally distributed were log transformed to ameliorate a positive skew^32^ or had outliers removed. We failed to asked participants about gender identity, which limits the generalizability of our research.

The procedures, hypotheses, and pre-analysis plan were pre-registered and data are publicly available in the Open Science Framework Repository^38^.

## 3. Results

### 3.1 Changes in dietary behaviors

Significantly more participants prefer salty foods after compared to before losing their sense of smell (X^2^=6.3, df=1, p=0.01; figure 1c), and significantly fewer participants prefer fatty foods after compared to before losing their sense of smell (X^2^=4.5, df=1, p=0.03; figure 1e), but there were no differences in preferences for sweet (X^2^=2.3, df=1, p=0.13; figure 1a), sour (X^2=^1.3, df=1, p=0.3; figure 1b), bitter (X^2^=0.5, df=1, p=0.5; figure 1d), or spicy tastes (X^2^=0.6, df=1, p=0.5; figure 1f). Forty-eight percent of participants indicated that they gained weight, while 30% indicated that they lost weight since the onset of their smell disorder. Seventy percent of participants indicated that they use more herbs and spices, and 58% of participants report eating more spicy foods today compared to before losing their smell, while 63% of participants reported eating more salt now compared to before their smell loss (Table 2). Responses to all questions can be found in Table 2. There were no differences between smell disorders for any questions.

**Figure 1.**
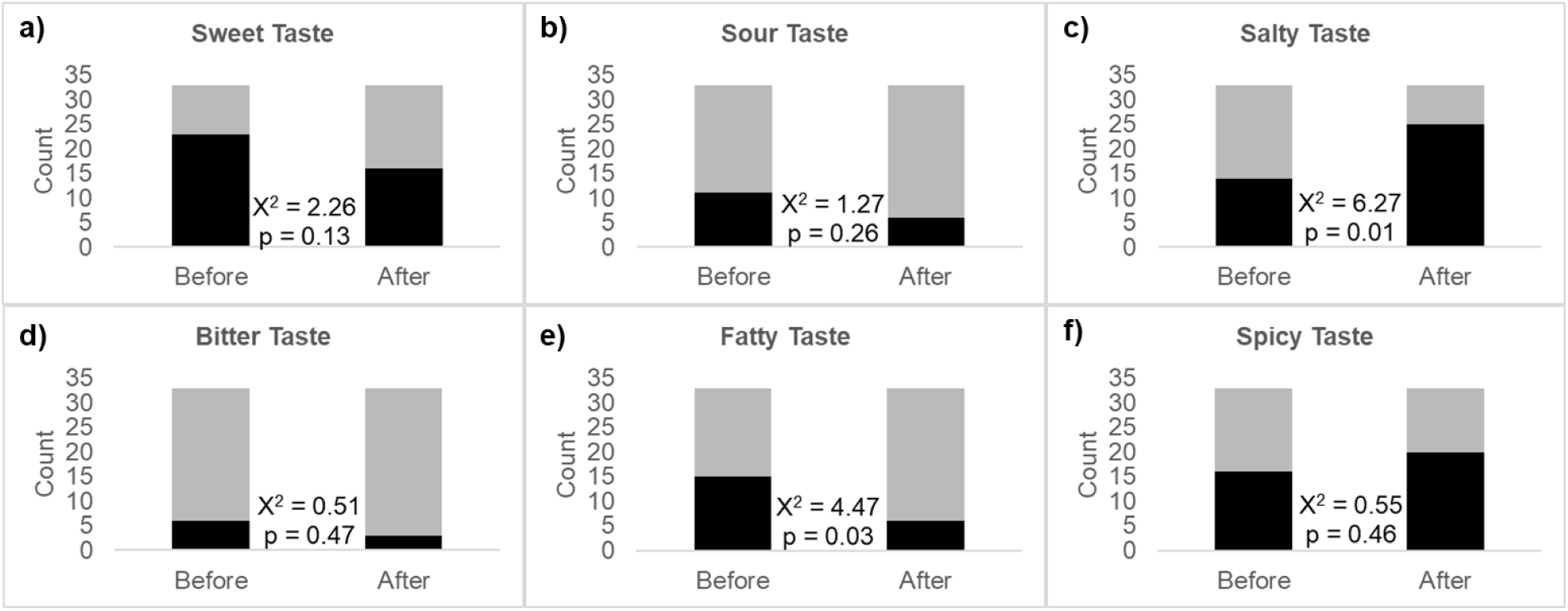
Changes in taste preferences before and after smell loss. Black represents those who prefer that taste, gray represents those who do not prefer that taste. n=33.

**Table 2.** Changes in dietary behaviors questionnaire. **n=33.**

| Question | More, n (%) | Less, n (%) | No Change, n (%) |
| --- | --- | --- | --- |
| Do you eat more or less now? | 10 (30) | 12 (36) | 11 (33) |
| Do you eat on a more or less regular schedule? | 4 (12) | 14 (42) | 15 (45) |
| Do you use more or less spices today? | 23 (70) | 7 (21) | 3 (9) |
| Do you invite people for dinner more or less frequently? | 1 (3) | 19 (58) | 13 (39) |
| Are you looking more or forward to dinner invitations? | 2 (6) | 18 (55) | 13 (39) |
| Do you need more or less time to eat a meal? | 3 (9) | 7 (21) | 23 (70) |
| Do you go out to eat to a restaurant more or less frequently? | 1 (3) | 19 (58) | 13 (39) |
| Do you eat more or less meals a day now? | 1 (3) | 17 (52) | 15 (45) |
| Do you eat more or less in between meals now? | 8 (24) | 12 (36) | 13 (39) |
| Do you spend more or less money on food now? | 11 (33) | 10 (30) | 12 (36) |
| Do you pay more or less attention to low caloric nutrition? | 3 (9) | 11 (33) | 19 (58) |
| Do you eat more or less cake today? | 7 (21) | 17 (52) | 9 (27) |
| Do you pay more or less attention to healthy nutrition now? | 7 (21) | 12 (36) | 14 (42) |
| Do you eat more or less sweets today? | 12 (36) | 18 (55) | 2 (9) |
| Do you eat more or less yogurt/milk products now? | 12 (36) | 9 (27) | 12 (36) |
| Do you eat more or less cheese now? | 12 (36) | 9 (27) | 12 (36) |
| Do you eat more or less fruits now? | 12 (36) | 13 (39) | 8 (24) |
| Do you eat more or less vegetables and salad now? | 9 (27) | 17 (52) | 7 (21) |
| Do you eat more or less spicy foods now? | 19 (58) | 5 (15) | 9 (27) |
| Do you eat more or less low-fat food now? | 4 (12) | 7 (21) | 22 (67) |
| Do you eat more or less high-fat food now? | 12 (36) | 7 (21) | 14 (42) |
| Do you use more or less artificial sweeteners now? | 4 (12) | 10 (30) | 19 (58) |
| Do you use more or less sugar now? | 6 (18) | 12 (36) | 15 (45) |
| Do you use more or less salt now? | 21 (63) | 4 (12) | 8 (24) |
| Do you drink more or less coffee or tea today? | 11 (33) | 6 (18) | 16 (48) |
| Do you drink more or less alcoholic beverages today? | 2 (6) | 15 (45) | 16 (48) |
| Do you drink more or less sweet drinks today? | 7 (21) | 17 (52) | 9 (27) |
| Is your daily intake of liquids higher or lower today? | 12 (36) | 4 (12) | 17 (52) |

## 3.2 24-hour urinary sodium

The average sodium intake in all participants was 2992 ± 279 mg/day (n=25). Since men and women have different levels of sodium intake, and there were only four males in this study, we only analyzed sodium intake in women. The average sodium intake for women included in this study was 2893 ± 258 mg/day (n=22). This is not significantly different compared to previous population estimates in women (3039 ± 99 mg/day; t=0.3, df=426, p=0.3), however, it is still higher than the recommended levels of sodium intake per day in the United States (2300 mg/day). Average sodium intake was not significantly different between smell disorders (supplementary table 1; F-value=0.7, df=2, p=0.5).

### 3.3 Moderate amounts of capsaicin increased salt taste intensity in model tomato soup samples

There was a significant effect of capsaicin (F_2,568_=165, p<0.001), where participants rated the soups with low (p<0.001) and moderate (p<0.001) amounts of capsaicin spicier than the soups with no capsaicin, and the soups with moderate amounts of capsaicin were rated as spicier than the soups with low amounts of capsaicin (p<0.001), indicating that participants could perceive the three different capsaicin levels, which was expected (Figure 2a). There was also an effect of objective disorder (F_1,19_=8.4, p=0.009), where those with anosmia (30.6 ± 2.1) rated the soups twice as spicy as participants with hyposmia (14.5 ± 1.1).

**Figure 2.**
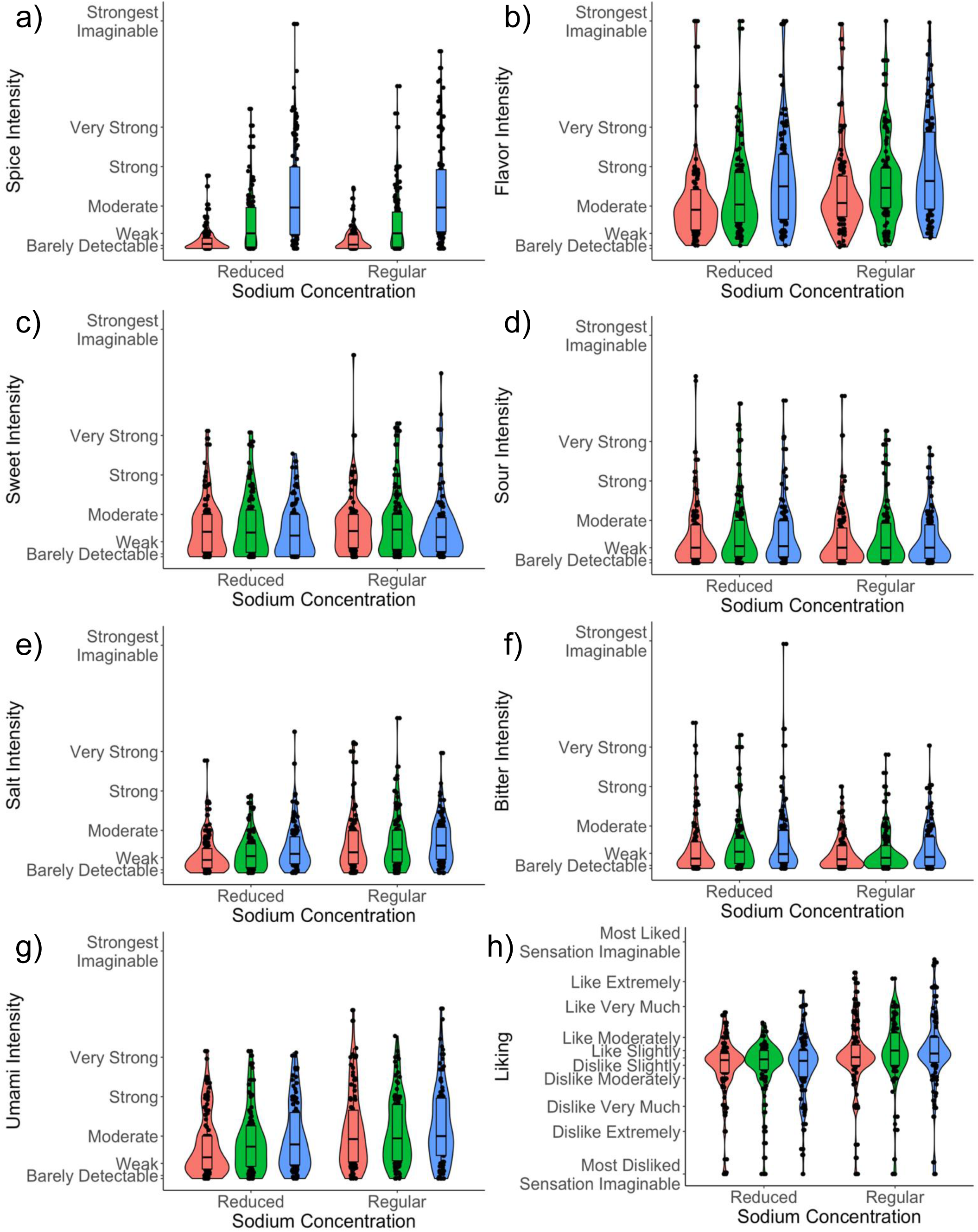
Intensity ratings for a) spice, b) flavor, c) sweet, d) sour, e) salt, f) bitter, g) umami, and h) liking ratings for soups with two sodium concentrations and three capsaicin concentrations. Pink bar = no capsaicin, green bar = low capsaicin, blue bar = moderate capsaicin. n=29.

There was a significant effect of sodium concentration on taste quality intensities, such that the regular sodium soup was rated more sweet (F_1,626_=6.4, p=0.01; Figure 2c), more salty (F_1,570_=32.1, p<0.001; Figure 2e), less bitter (F_1,570_=13.3, p<0.001; Figure 2f), and more umami (F_1,570_=35.1, p<0.001; Figure 2g) than the reduced sodium soup. There was also a significant effect of capsaicin level on umami (F_2,570_=5.6, p=0.004), sweet (F_2, 626_=3.3, p=0.04), bitter (F_2,570_=5.4, p=0.005), and salt (F_2,570_=5.5, p=0.004) taste intensities, such that the soups with moderate amounts of capsaicin were more umami (Figure 2g; p<0.001), less sweet (p=0.02, Figure 2c) and more bitter (Figure 2f; p<0.001) than soups with no capsaicin added. Soups with moderate (p<0.001) and low (p=0.02) capsaicin were also more salty than soups with no capsaicin (Figure 2e). There were no significant effects of sodium level or capsaicin levels on sour intensity ratings (F_2,570_=1.1, p=0.3; Figure 2d). There was also a significant sodium (F_1, 570_=14.4, p<0.001) and capsaicin (F_2,570_=27.3, p<0.001) effect on flavor intensity, where regular sodium soups were rated more flavorful than the reduced sodium soups (p<0.001), and soups with moderate (p<0.001) and low (p<0.001) amounts of capsaicin were more flavorful than soups with no capsaicin added, and soups with moderate (p<0.001) capsaicin added were more flavorful than soups with low capsaicin added (Figure 2b). Regular sodium soups were liked more than the reduced sodium soups (F_1, 569_=35.5, p<0.001), but there was no effect of capsaicin level on liking (F_2,568_=1.8, p=0.1; Figure 2h). Thus, adding moderate amounts of capsaicin made the soups more flavorful, salty, bitter, and umami, and less sweet, compared to the soup when no capsaicin was added, but this did not translate to the soups being more liked.

### 3.4 Adding spice to model foods increased liking in a food-specific manner

There was a significant food*spice level interaction effect on spice intensity (F_2, 603_=5.3, p=0.005), where both pasta and chocolate food samples with low (p<0.001 for both) and moderate (p<0.001 for both) amounts of spice were rated more spicy than the pasta and chocolate with no spice added, and the pasta and chocolate samples with moderate (p<0.001 for both) amounts of spice added were rated more spicy than pasta and chocolate samples with low amounts of spice added, respectively. Thus, participants were able to perceive differences between the three spice levels (Figure 3a).

**Figure 3.**
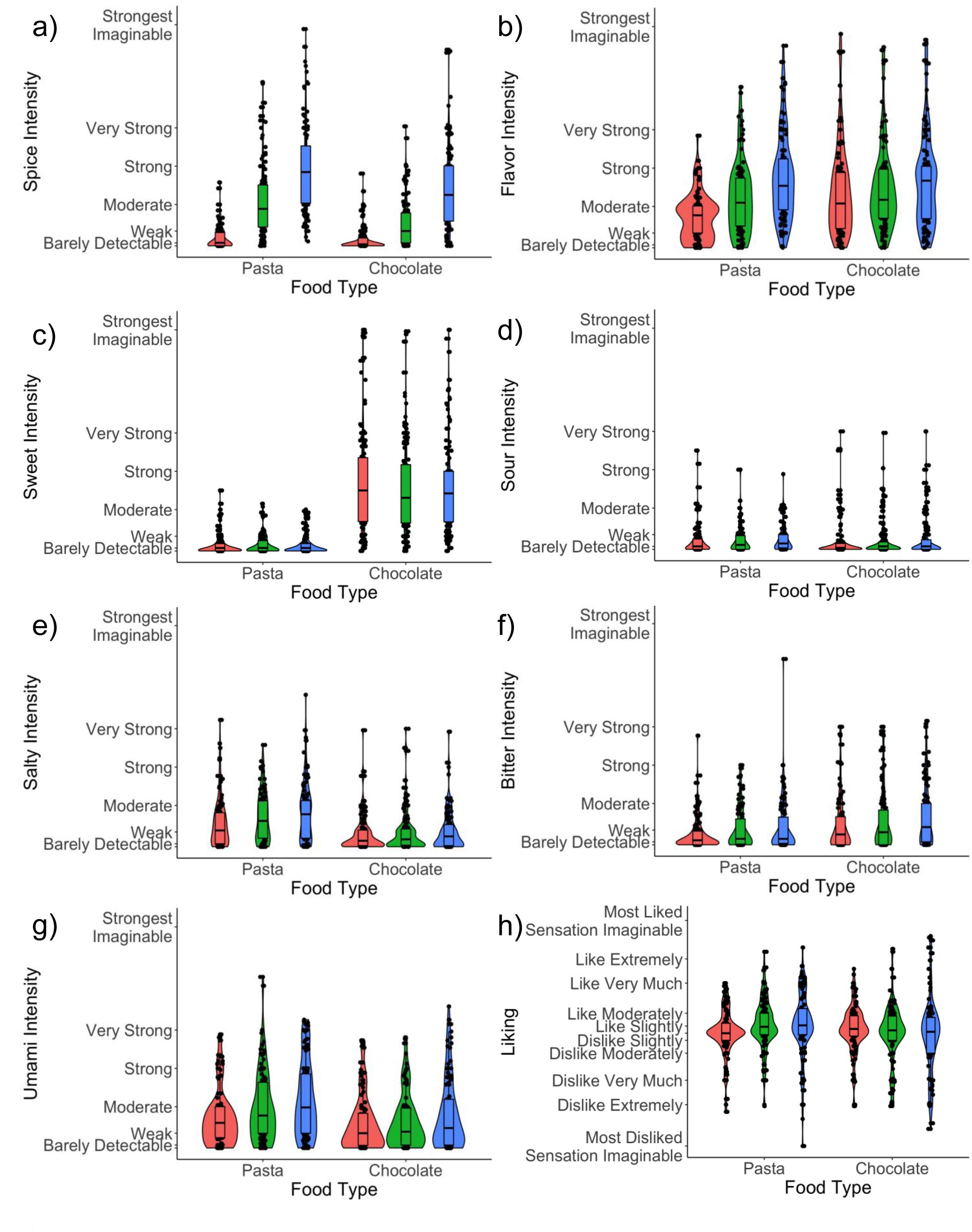
Intensity ratings for a) spice, b) flavor, c) sweet, d) sour, e) salt, f) bitter, g) umami, and h) liking ratings for food samples (pasta and chocolate) with three spice levels added. Pink bar = no spice, green bar = low spice, blue bar = moderate spice. Data are means ± SE. n=31.

There was a significant food*spice level interaction effect on total flavor intensity (F_2,581_=14.1, p<0.001; Figure 3b), where pasta samples with low (p<0.001) and moderate (p<0.001) amounts of spice were more flavorful than pasta samples when no spice was added, and pasta samples with moderate (p<0.001) amounts of spice were more flavorful than pasta samples with low amounts of spice added. Chocolate samples with moderate amounts of spice were rated more flavorful than chocolate samples with no spice added (p=0.01; Figure 3b). Pasta samples were rated less sweet (F_1, 604_=1520.1, p<0.001; Figure 3c), more sour (F_1, 526_=20.5, p<0.001; Figure 3d), more salty (F_1,604_=184.3, p<0.001; Figure 3e), less bitter (F_1,603_=34.2, p<0.001; Figure 3f), and more umami (F_1,604_=75.6, p<0.001; Figure 3g) than chocolate samples. There was a significant effect of spice level on sour ratings (F_2, 524_=4.9, p=0.008), where food samples with moderate amounts of spice were rated more sour (p=0.004; Figure 3d) than food samples with no spice, but there was no significant interaction between food type and spice level (F_2, 524_=0.06, p=0.9). There were also significant effects of spice level on salt (F_2,603_=6.4, p=0.002), bitter (F_2,602_=10.4, p<0.001), and umami (F_2, 603_=12.2, p<0.001) ratings, where food samples with low and moderate amounts of spice added were rated more salty (p=0.04 and p<0.001, respectively; Figure 3e), bitter (p=0.01 and p<0.001, respectively; Figure 3f) and umami (p=0.02 and p<0.001, respectively; Figure 3g) than food samples with no spice, and moderate amounts of spice added were rated more umami than food samples with low spice (p=0.04), but there was no significant interaction between food type and spice level. Finally, pasta samples were liked more than chocolate samples (F1,538=5.8, p=0.02; Figure 3h) and there was a significant food*spice level interaction effect (F_2,530_=6.3, p=0.002), where adding a moderate amount of spice to pasta samples increased liking compared to pasta samples with no spice added (p<0.001). Furthermore, younger participants (F_1, 22_=4.9, p=0.04), men (F_1,24_=5.0, p=0.04), and those with anosmia (F_1,21_=6.0, p=0.02) had higher liking ratings compared to older participants, females, and those with hyposmia.

## 4. Discussion

Flavor perception and food liking are negatively affected when someone loses their smell, leading to compensatory behaviors, such as increased salt use. The purpose of this study was to establish sodium intake and eating behavior changes in those with smell loss, quantify liking of a meal with and without capsaicin in individuals with smell loss, and determine whether adding capsaicin increases salt taste intensity in individuals with smell loss.

The majority of participants self-reported that they consumed more herbs and spices (70%), spicy foods (58%) and salt (63%) after losing their sense of smell. Participants also preferred salty tastes more after losing their sense of smell. These results are consistent with other studies and anecdotal reports of participants altering their diets to compensate for the lack of flavor, specifically by adding in tastes (like salt)^11,14^ and enhancing the chemesthetic aspect of flavor (spices)^10,11,15,16^, and preferring salty foods^10^. Preferring sweet tastes is also frequently reported in those with smell loss^11^, but there were no significant differences in reported preference for sweet tastes before and after smell loss in this study, and the majority of our participants indicated no change in their sugar consumption, or reported consuming less sugar after smell loss.

Despite participants self-reporting a higher salt intake and salt taste preferences, sodium intake measured by 24-hour urine was not significantly higher compared to population averages. However, these results should be interpreted with caution, since we do not know what their sodium intake was before losing their sense of smell and therefore do not know if, or in which direction, it changed. If the majority of salt added to the diet is from discretionary sources, which would provide the most direct taste, there would likely only be a small effect on sodium intake^39^. Excess sodium intake in the population is largely driven by consumption of processed foods, which have a high sodium content, but do not necessarily taste salty^39^. Future studies should assess which food sources are contributing the most sodium to the diet in people with smell loss.

Pasta samples were liked more with moderate amounts of spice added, but liking of tomato soup or chocolate did not change with the addition of spiciness. People who like spicy foods may also be more receptive to the samples with moderate spice added, but there was no effect of spice intake frequency on results in this study. Pasta samples may have been the only food that increased in liking because adding spice to pasta is more common than tomato soup or chocolate, which may have been more of a novel flavor. Previous studies report that individuals with smell loss are less attracted to novel foods than people with a normal sense of smell^40^, thus our participants may have been more wary of these samples. Exposure to novel foods multiple times may be needed to see increases in liking in individuals with smell loss. Furthermore, several of our participants indicated that they have parosmia, and may dislike all foods, even if a palatable taste is added. Nevertheless, our results still provide evidence that adding low and moderate amounts of spiciness to food may help increase how much a food is liked, which can promote intake of that food. Future studies should assess whether this can be used as a strategy to improve diet quality by increasing vegetable intake in individuals with smell loss, which is lower than the population with a normal sense of smell^41^, and whether these sensory strategies work in some people more than others.

Participants reported that soup samples with low to moderate amounts of capsaicin were more salty compared to soup samples without capsaicin added. Our results support a growing body of evidence that capsaicin increases salt taste intensity, particularly at low to moderate concentrations of salt^24,25,42–45^. This suggests that capsaicin may be particularly beneficial in increasing perceived saltiness of reduced sodium food products, which could aid in their acceptance. However, the salt taste enhancement was weak in our study, and cross-modal interactions between taste, smell, and chemesthesis may be necessary to see a stronger enhancement of salt taste^25,42^. It is noteworthy that other studies have been conducted in Asian countries, where spice consumption is high. To our knowledge, this is the first study that has been done in a US sample, and in those with smell loss.

While the mechanism for the enhancement of salt taste by capsaicin is unknown, it could be that a TRPV1 (the receptor for capsaicin) variant plays a role in salt taste detection^46,47^.

While there is growing evidence supporting capsaicin increasing salt taste intensity, whether this actually helps reduce salt intake over time has not been tested. Previous studies have modeled sodium reduction with salt taste intensity enhancement, and have found that a 51% enhancement in salt taste intensity would result in about a 10% reduction in sodium^25^. Another study found that sodium reduction was dependent on the sensitivity of participants to salt, but that 0.41 g/L Sichuan pepper oleoresin (which includes trigeminal and odor component) could increase salt taste intensity and reduce sodium at levels upwards of 3.93 points on a gLMS and 39%, respectively, for the semi-sensitive group, and 3.27 points on a gLMS and 39%, respectively, for the hypersensitive group^42^.

Despite many studies reporting that capsaicin increases salt taste intensity, other studies have found that capsaicin reduces salt taste intensities^48,49^. However, the reduction in taste intensity occurs when the burning sensation is particularly high, either from high levels of capsaicin used or in infrequent spice consumers who are more sensitive to the burn. Thus, there is a cutoff where capsaicin either enhances tastes or diminishes tastes. To use capsaicin as a flavor enhancement strategy, low to moderate levels should be used.

One limitation of this study is the lack of a normosmic control group. However, the purpose of this study was specifically to determine if capsaicin increases food liking and salt taste intensity in those with smell loss. Nonetheless, a normosmic control would help to understand if the salt taste enhancement is of the same magnitude in those with smell loss, and could have given a better comparison for sodium intake. Another limitation is that we were not able to collect 24-hour urine sodium intake from all participants due to travel issues and unwillingness to collect the sample, resulting in a sample size of only 22 female participants. Furthermore, we did not measure creatinine to assess completeness of the sample, therefore we do not know the accuracy of the sodium intake measures. However, only participants who had urine volumes greater than 500 mL were included, which has been used in previous studies as a guide to determine whether a sample is complete or not^35^.

This is one of the first studies to assess a sensory strategy to improve food liking in a way that promotes diet quality in people with smell loss. There is a need for more sensory nutrition research in this growing population with smell loss; they are a growing population which suffers from lack of food enjoyment, and is at risk for worse diet quality, a higher BMI and waist circumference, and more chronic diseases than those with a normal sense of smell^41^. More sensory nutrition research is needed into this growing population with smell loss in order to improve diet quality, eating enjoyment, and health, which will likely result in improvements to their quality of life^23^.

## 5. Conclusions

People who lose their sense of smell struggle with reduced flavor perception and eating enjoyment, resulting in compensatory strategies that can have a negative effect on health if maintained long-term. In this study, we found that participants with smell loss reported consuming more herbs and spices, salt, and spicy foods. They also consume more sodium than is recommended in the United States, but not more than population averages with normosmia. Capsaicin increases total flavor and salt taste intensity in people with smell loss. Adding spice to foods can help increase food liking, but this is dependent on the food to which the spice is added. Thus, adding spice to foods is a useful strategy to help improve food liking, and flavor and salt taste intensity. Future studies should assess whether this strategy can be used to reduce sodium intake and improve diet quality.

### Abbreviations

gLMS: general labeled magnitude scale
LHS: labeled hedonic scale

## Data Availability

All data produced in the present study will be available online at Open Science Framework Repository upon publication of the manuscript.

https://doi.org/10.17605/OSF.IO/WP4GM

## 6. Acknowledgements

This work would not be possible without people who are willing to participate in the research. We are grateful to all participants in this study, as well as the AbScent and STANA communities for allowing us to recruit from their communities.

## 7. Author Contributions

SRH: conceptualization, methodology, formal analysis, investigation, visualization, supervision, project administration, funding acquisition, writing – original draft, writing – review & editing. CB: investigation, writing – review & editing. PHD: conceptualization, methodology, supervision, writing – review & editing.

## Supplementary Information

**Supplementary Table 1.** 24-hour urine sodium results in female participants only.

| **Group** | **n** | **24-hour Urinary Sodium Excretion (mg)** | **F-value (p-value)** |
| --- | --- | --- | --- |
| Overall | 22 | 2893 ± 258 | 0.67 (p=0.526) |
| Anosmia | 5 | 2336 ± 337 |  |
| Hyposmia | 8 | 3043 ± 517 |  |
| Hyposmia + Parosmia | 9 | 3067 ± 399 |  |

Data are means ± SE.

**Supplementary Table 2.**
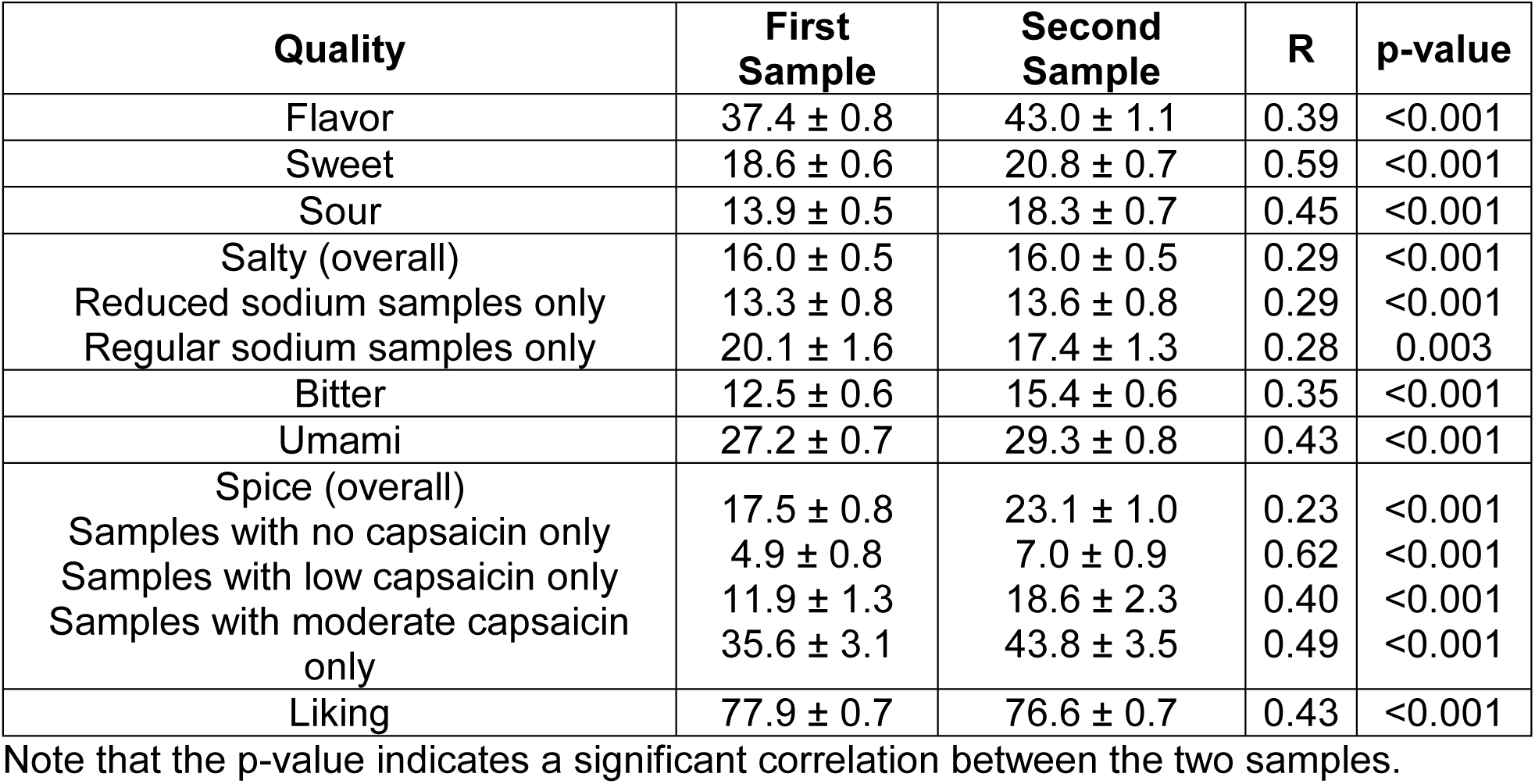
Within-session test-retest reliability for model soup samples.

**Supplementary Table 3.**
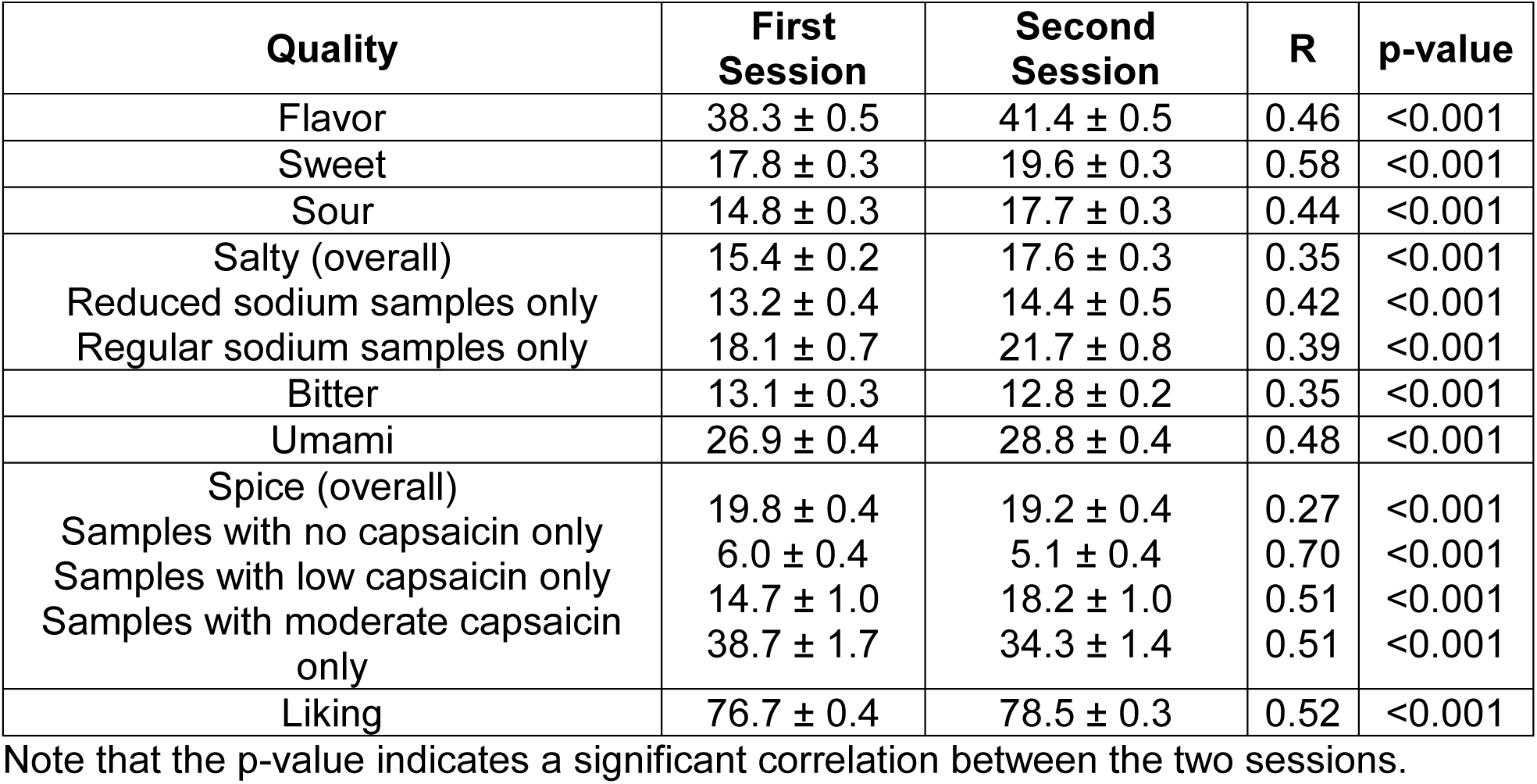
Between-session test-retest reliability for model soup samples.

**Supplementary Table 4.** Within-session test-retest reliability for model food samples.

| Quality | First Sample | Second Sample | R | p-value |
| --- | --- | --- | --- | --- |
| Flavor | 35.8 ± 0.8 | 37.1 ± 0.8 | 0.47 | <0.001 |
| Sweet | 25.9 ± 1.0 | 24.6 ± 0.9 | 0.23 | <0.001 |
| Sour | 6.0 ± 0.3 | 5.6 ± 0.3 | 0.42 | <0.001 |
| Salty | 14.6 ± 0.5 | 13.1 ± 0.4 | 0.34 | <0.001 |
| Bitter | 11.3 ± 0.5 | 13.3 ± 0.5 | 0.42 | <0.001 |
| Umami | 21.1 ± 0.7 | 21.1 ± 0.6 | 0.46 | <0.001 |
| Spice | 22.9 ± 0.8 | 23.3 ± 0.8 | 0.16 | <0.001 |
| Samples with no spice only | 3.9 ± 0.3 | 5.6 ± 0.4 | 0.40 | <0.001 |
| Samples with low spice only | 21.5 ± 1.1 | 23.7 ± 1.0 | 0.19 | <0.001 |
| Samples with moderate spice only | 46.3 ± 1.5 | 46.7 ± 1.4 | 0.46 | <0.001 |
| Liking | 83.1 ± 0.7 | 80.8 ± 0.6 | 0.39 | <0.001 |

Note that the p-value indicates a significant correlation between the two samples.

**Supplementary Table 5.** Between-session test-retest reliability for model food samples.

| Quality | First Sample | Second Sample | R | p-value |
| --- | --- | --- | --- | --- |
| Flavor | 37.1 ± 0.5 | 38.4 ± 0.4 | 0.45 | <0.001 |
| Sweet | 25.6 ± 0.5 | 24.3 ± 0.5 | 0.24 | <0.001 |
| Sour | 6.1 ± 0.2 | 6.9 ± 0.2 | 0.46 | <0.001 |
| Salty | 13.6 ± 0.3 | 14.7 ± 0.3 | 0.3 | <0.001 |
| Bitter | 13.2 ± 0.3 | 13.4 ± 0.3 | 0.46 | <0.001 |
| Umami | 22.6 ± 0.4 | 23.7 ± 0.4 | 0.46 | <0.001 |
| Spice | 23.9 ± 0.5 | 25.4 ± 0.4 | 0.17 | <0.001 |
| Samples with no spice only | 3.6 ± 0.5 | 4.2 ± 0.6 | 0.45 | <0.001 |
| Samples with low spice only | 22.4 ± 2.2 | 19.6 ± 1.9 | 0.21 | 0.015 |
| Samples with moderate spice only | 42.7 ± 2.7 | 46.0 ± 2.7 | 0.42 | <0.001 |
| Liking | 81.0 ± 0.3 | 81.1 ± 0.3 | 0.32 | <0.001 |

Note that the p-value indicates a significant correlation between the two sessions.

